# An Agent Based Model for assessing spread and health systems burden for COVID-19 in Rangareddy district, Telangana state, India

**DOI:** 10.1101/2020.06.04.20121848

**Authors:** Narassima M S, Guru Rajesh Jammy, Rashmi Pant, Lincoln Choudhury, Aadharsh R, Vijay Yeldandi, Anbuudayasankar S P, Rangasami P, Denny John

## Abstract

**Objectives:** To evaluate the transmission dynamics and health systems burden of COVID-19 with and without interventions, using an Agent Based Modeling (ABM) approach on a localized synthetic population.

**Study design:** A synthetic population of Rangareddy district, Telangana state, India, with 5,48,323 agents and simulated using an ABM approach for three different scenarios.

**Methods:** The patterns and trends of the COVID-19 in terms of infected, admitted, critical cases requiring intensive care and/ or ventilator support, mortality and recovery were examined. The model was simulated over a period of 365 days for a no lockdown scenario and two Non-Pharmaceutical Intervention (NPI) scenarios i.e., 50% lockdown and 75% lockdown scenarios. Sensitivity Analysis was performed to compare the effect of change of parameters on disease dynamics.

**Results:** Results revealed that the peak values and slope of the curve declined as NPI became more stringent. The peak values could facilitate policymakers to plan the required capacity to accommodate influx of hospitalizations.

**Conclusions:** ABM provides better insight into projections compared to compartmental models. The results could provide a platform for researchers and modelers to explore using ABM approach for COVID-19 projections with inclusion of interventions and health system preparedness.

## 1. Introduction

The first reported case of the novel coronavirus (COVID-19 or SARS-CoV-2) in India dates back to January 30, 2020 when it was also announced as pandemic by WHO [52; 78]. Since then, epidemic has spread across India infecting 44,94,389 people with 9,27,545 active cases, 34,90,908 recovered cases and 75,328 deaths as on Sep 10, 2020 [17]. Globally, COVID-19 has spread across 213 nations, infecting 28,035,700 people worldwide and claiming 908,991 lives as on Sep 10, 2020, posing a global health emergency [18; 42; 75].

In a country like India having a denser population, the situation poses a serious challenge [37]. There are several underlying factors such as age, comorbidities, exposure to air pollution, amount of exposure to virus, etc., that determine transmission dynamics [43]. Travel restrictions and [15; 44] other Non-Pharmaceutical Interventions (NPIs) such as restrictions on public gatherings, intra-city movements, etc., may flatten the curve. It is important to understand the time required between exposure and complete recovery, using these interventions, to take timely responsive actions against COVID-19 [80].

Modelling is an effective tool to provide real-time projections and obviates the burden of investing time, cost and risks [48]. Several models on COVID-19 projections exist and most of them are compartmental models. Since compartmental models assume homogeneity in the compartments they do not account for individual level variations and interactions within the system, Agent Based Modeling (ABM) is a tool which considers these factors and may provide better insights [26]. ABM considers the interaction between agents whilst also distinguishing them based on their individual parameters [12; 71]. Advancements in computational capabilities have increased interest among researchers across various verticals, including public health in recent times on ABM [33; 31; 32].

ABM incorporates actions of agents within the system, helping the model comprehend infection spread dynamics better [5; 60]. ABM follows a bottom-up approach wherein result of behaviour of individuals, defined as agents within the system [13; 27]. ABM allows to define unique characteristics to the agents to make each of them behave distinctly [13]. Mixing patterns among the agents within a system play a vital role in dynamic transmission models for close contact infections [8].

In the past, ABMs have been employed to address various infectious diseases such as, a bioterrorist introduction of smallpox [34], design of vaccination strategies for influenza [16], curtail transmission of measles through contact tracing and quarantine [23], control of tuberculosis [54], implementation of distancing measures and antiviral prophylaxis to control H5N1 influenza A (bird flu) [25] and devise evacuation strategies in the event of airborne contamination [24].

An evaluation of use of ABM on COVID-19 globally suggests its use to measure the effects of lockdown on transmission dynamics [39; 50; 69; 72], post-lockdown measures [36], use of control measures (face mask, social distancing) [36; 41], isolation of vulnerable proportion of population [36; 35], contact tracing, intelligence of agents (based on awareness level [69] or protection level), contact tracing measures [41; 69], good practices such as sneezing into one’s hands [39], both direct (upon contact) and indirect transmission (through suspended particles) [39], scheduled-based contacts [20; 36; 39] with close circle and in work, transport and public places [35; 39; 51], viral-load based transmissibility [41], examination of genomic sequencing to determine the spread [66], etc.

In India, several COVID-19 models have been conducted based on Susceptible (S), Exposed (E), Infective (I) and Recovered (R) (SEIR) [14; 67; 74; 79], Susceptible (S), Exposed (E), Symptomatic (I), Purely Asymptomatic (P), Hospitalized or Quarantined (H), Recovered (R) and Deceased (D) (SIPHERD) [49], mathematical models [2; 68], etc., to compare the spread during lockdown and no lockdown scenarios. Nation-wide models restrict the policymakers locally to devise strategies based on the results as they might not fit properly to the locality [10]. In countries like India where people are diverse in all respects like population dynamics, contact network, migrating population, nature of work, etc., local models might prove effective and would assist policymakers to take local decisions for disease mitigation. The present study aims at an ABM to examine the patterns and trends of spread and the effect of NPIs in a synthetic population of a region in Telangana state of India.

## 2. Methods

### 2.1. Study Design

A synthetic population of Telangana state, India, has been developed, the details of it are presented elsewhere [62]. We used 5,48,323 agents from this population from the Rangareddy district. The modeling follows an ABM approach using AnyLogic 8.5.2 University edition to model the interaction environment [6]. The entire simulation and reporting follows the International Society for Pharmacoeconomics and Outcomes Research (ISPOR-SMDM) Modeling Good Research Practices and ethical good practice in modelling [9; 11; 65]. These guidelines were used so that the assumptions, scope and shortcomings of the model are transparent to the readers and policymakers.

### 2.2. Synthetic population

Synthetic population is one of the commonly used approaches to represent a group of people, preserving the confidentiality of individuals. Synthetic population has statistical equivalence with the original population being represented and is indistinguishable from the census data [1]. For this study, we used a synthetic population developed for Rangareddy, Telangana state consisting of 5,48,323 people (representing 10.35 % of Rangareddy’s population (n-52,96,741) as per Census of India 2011), to demonstrate the ABM [62; 63]. The population was categorized based on age as less than 5, 5 to 59, and above 60 with 47,039, 4,59,372 and 41,912 agents respectively [8; 43].

### 2.3. Transmission rates of COVID-19

Based on the WHO report on COVID-19 (16 to 24 Feb 2020), the transmission rates were set to vary from 1 to 10 percent [4].

### 2.4. Contact network

The contact network plays a vital role in transmission dynamics. For contact rate estimation, we used a study from Ballabgarh, India, which determined the contact rates for close contact infections [46]. The dataset representing the number of people met by each individual was input into the ‘Input Analyzer’ tool of Arena (Version 16.00.00002). Input Analyzer provides fitting distributions with associated errors of fitting. The contact rates of each group were found to follow normal distributions using “Input Analyzer” (Table 1).

**Table 1:** Contact rate distribution of Ballabgarh [46]

| Years | Number of respondents | Average no. of people met per day | Min | Max | Distribution of contact rate(s) | Square Error |
| --- | --- | --- | --- | --- | --- | --- |
| <5 | 378 | 15.108 | 4 | 48 | NORM(15.1, 6.82) | 0.00656 |
| 5 to 59 | 2185 | 17.194 | 1 | 67 | NORM(17.2, 8.01) | 0.003202 |
| 60+ | 380 | 12.863 | 1 | 41 | NORM(12.9, 5.49) | 0.002328 |

The population densities of various townships of India were calculated based on Indian population proportion (table 2) [7; 57]. The population density of Ballabgarh was 551 people per square kilometer [46]. This was used to proportionately determine the contact rates of towns based on their respective population densities, assuming Density-Dependent (DD) contact rate [38; 58].

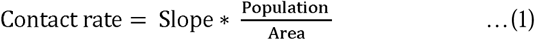

**Table 2:**
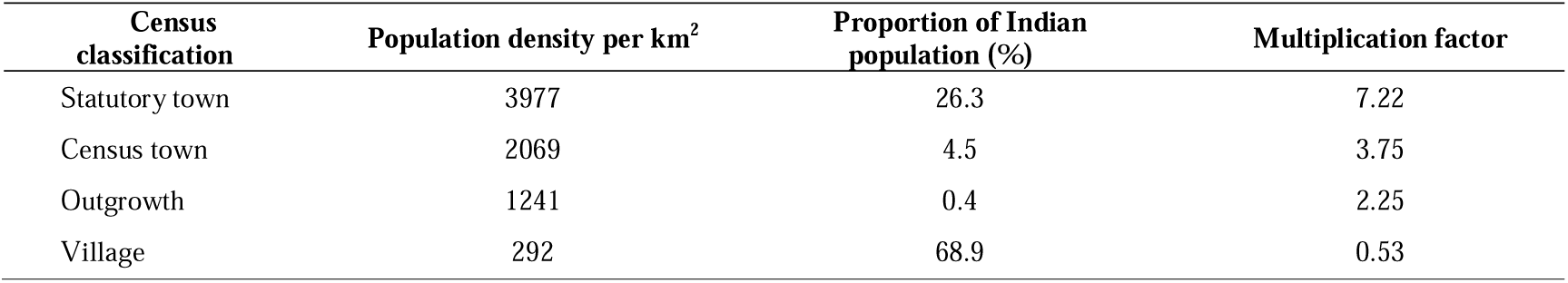
Classification of Indian towns with population proportions and densities [7]

For estimation of overall average contact rate, weights equal to the corresponding proportions of people living in different townships were multiplied to their corresponding multiplication factors (slope in equation (1)) (table 2). Contact rate was thus derived as the product of this slope and the ratio of the population density of a particular township to the population density of Ballabgarh.

The number of contacts made by people under each town category were multiplied by their respective multiplication factors as reported in Table 2. These values were integrated to Input Analyzer and their respective contact rate distributions were then determined. Contact rate for the present study followed Lognormal distributions (Lognormal(μ, σ, Min)) (table 3).

**Table 3:**
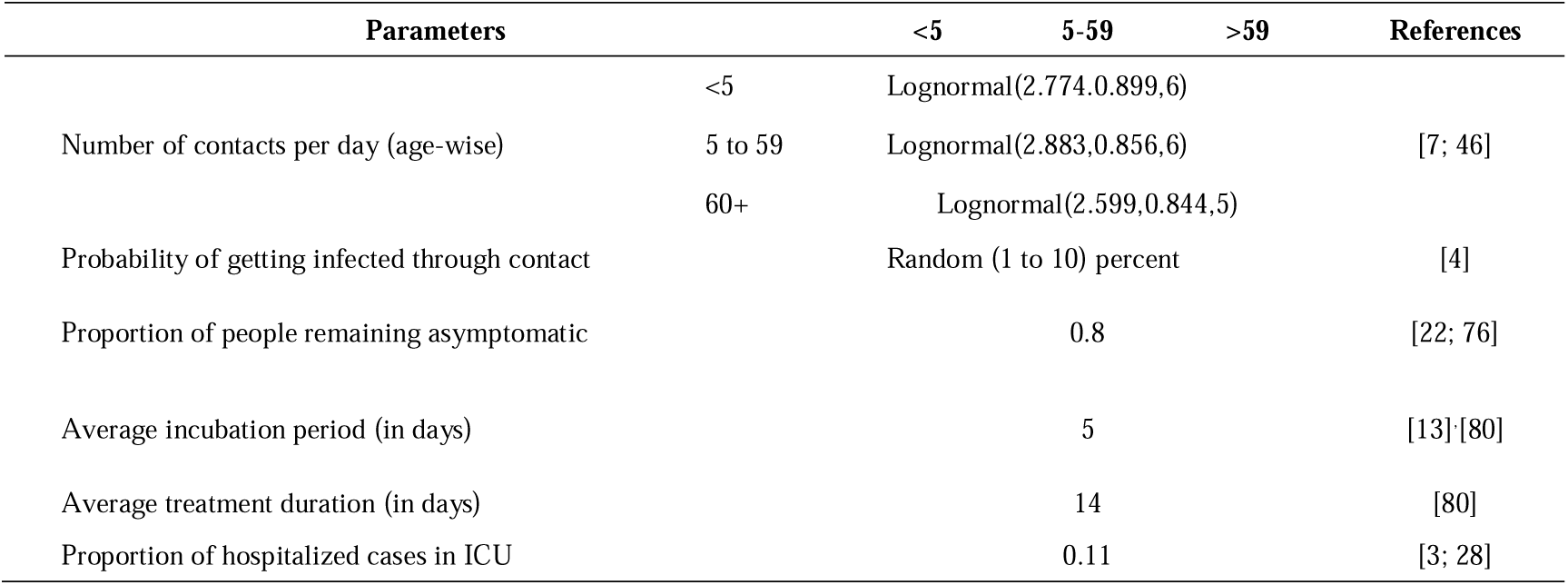
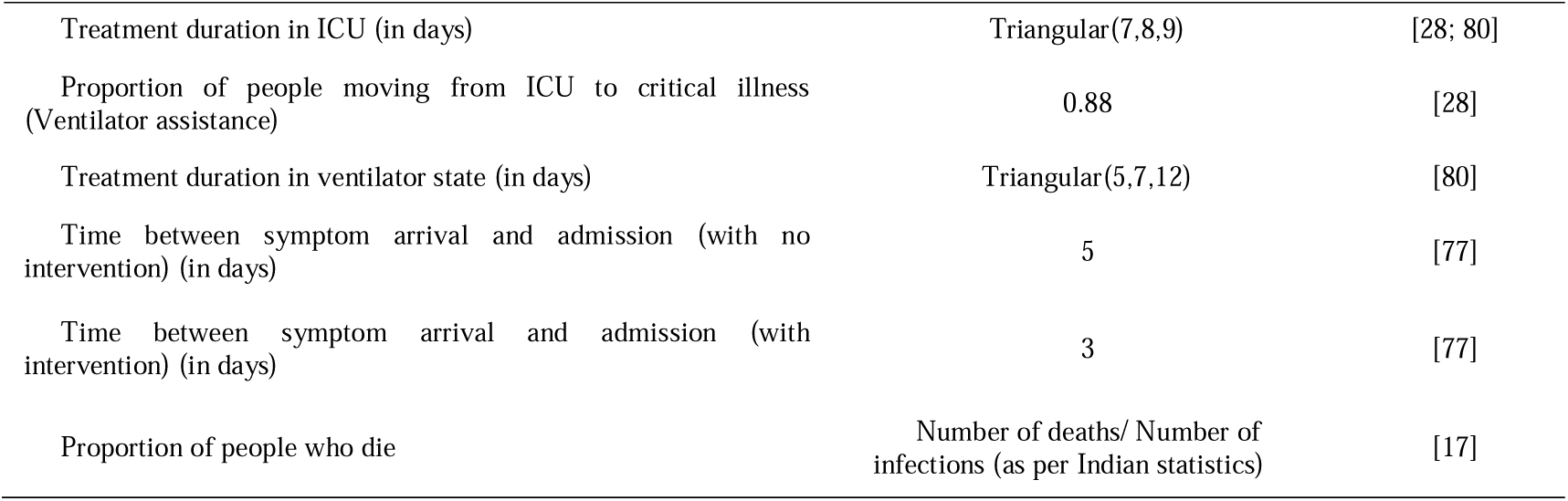
Parameters for the model

| Parameters |  | <5 | 5-59 | >59 | References |
| --- | --- | --- | --- | --- | --- |
| Number of contacts per day (age-wise) | <5 | Lognormal(2.774,0.899,6) |  |  | [7; 46] |
|  | 5 to 59 | Lognormal(2.883,0.856,6) |  |  |  |
|  | 60+ | Lognormal(2.599,0.844,5) |  |  |  |
| Probability of getting infected through contact |  | Random (1 to 10) percent |  |  | [4] |
| Proportion of people remaining asymptomatic |  |  | 0.8 |  | [22; 76] |
| Average incubation period (in days) |  |  | 5 |  | [13][80] |
| Average treatment duration (in days) |  |  | 14 |  | [80] |
| Proportion of hospitalized cases in ICU |  |  | 0.11 |  | [3; 28] |

|  |  |  |
| --- | --- | --- |
| Treatment duration in ICU (in days) | Triangular(7,8,9) | [28; 80] |
| Proportion of people moving from ICU to critical illness (Ventilator assistance) | 0.88 | [28] |
| Treatment duration in ventilator state (in days) | Triangular(5,7,12) | [80] |
| Time between symptom arrival and admission (with no intervention) (in days) | 5 | [77] |
| Time between symptom arrival and admission (with intervention) (in days) | 3 | [77] |
| Proportion of people who die | Number of deaths/ Number of infections (as per Indian statistics) | [17] |

### 2.5. State chart

A state chart represents the various states in which an agent would exist (figure 1) [30]. The initial state of all agents healthy, during the start of simulation. The agents would interact with other agents in the population and transmit the infection. Infected agents undergo an incubation period and turn out to be either symptomatic or asymptomatic. They continue to contact other agents and transmit till they get admitted or recover. Once admitted, agents undergo treatment and either decease or recover whilst in any of the three levels of infections represented by ‘admitted’, ‘ICU’ and ‘ventilator’ states [27].

**Figure 1:**
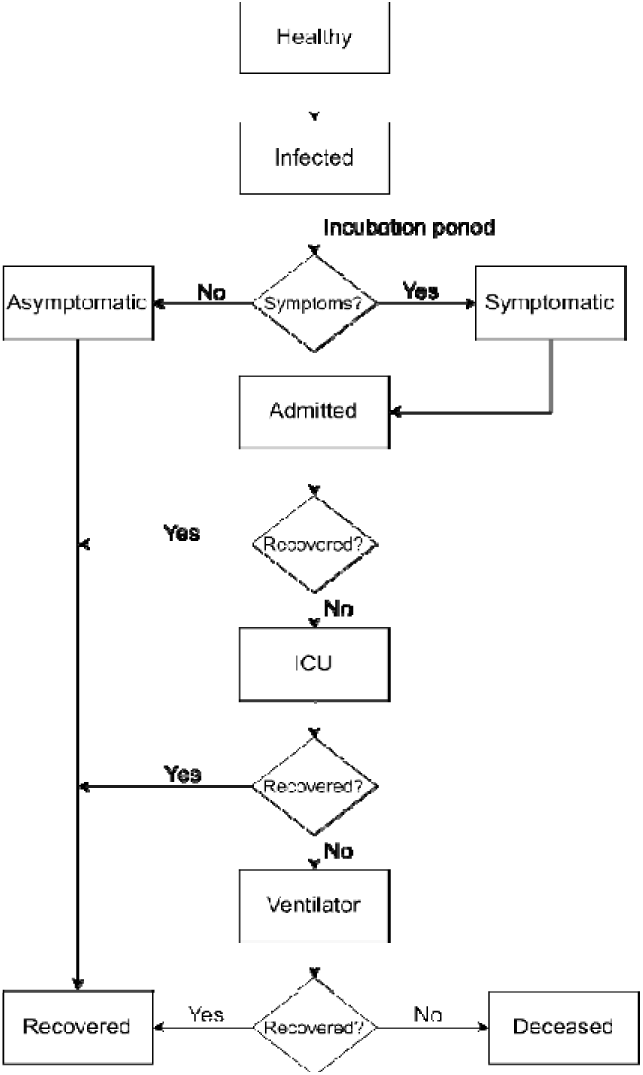
State chart for agent(s) (people)

### 2.6. Model parameters

Models of Infectious Disease Agent Study (MIDAS) has been used as a source of acquiring parameters through the pre-prints and manuscripts available [53]. Table 3 details the various parameters used for the model.

### 2.7. Model scenarios

In order to study the effect of minimization of number of contacts among the agents, three different scenarios were simulated for 365 days, results of which are discussed subsequently. First scenario, the “no lockdown” scenario includes no NPI put in place. In order to study the effect of lockdown, the contact rate of people needs to be reduced. To achieve this, the results of a study that presented the proportion of contacts made by individuals of different age groups were utilized. The number of contacts made at different locations namely home, school, work and others, as designated by the authors of the study was used to enact the lockdown [61]. To simulate the NPI scenarios, the number of people met in work and other places were reduced by 75 percent and 50 percent for the two scenarios whereas the contacts in school was completely discarded owing to the closure of schools. These resulted in reduced diffusion of the infection across the population, results of which are discussed subsequently.

## 3. Results

Simulations were run for different age groups as per the categorization for all the three scenarios. Detailed day-wise data of the number of people in each health state are provided in the supplementary files. The results of the same and their interpretations are discussed below.

It was observed that (Figure 2), number of uninfected people declines as the stringency of the imposed lockdown increases. After a duration of one year, proportion of people who remain uninfected was 28.53, 76.33 and 93.8 percent in No lockdown, 50% lockdown and 75% lockdown scenarios respectively. It was observed that (Figure 2 b) the spread of the infection observed as peak infections in scenarios 2 & 3 were 129779 and 33973 which would be reached in a period of just 33 and 25 days in a no lockdown condition. The peak values 191907, 37790 and 7986 in figure 2 c corresponds to 35%, 6.89% and 1.46% respectively of the initial uninfected population.

**Figure 2:**
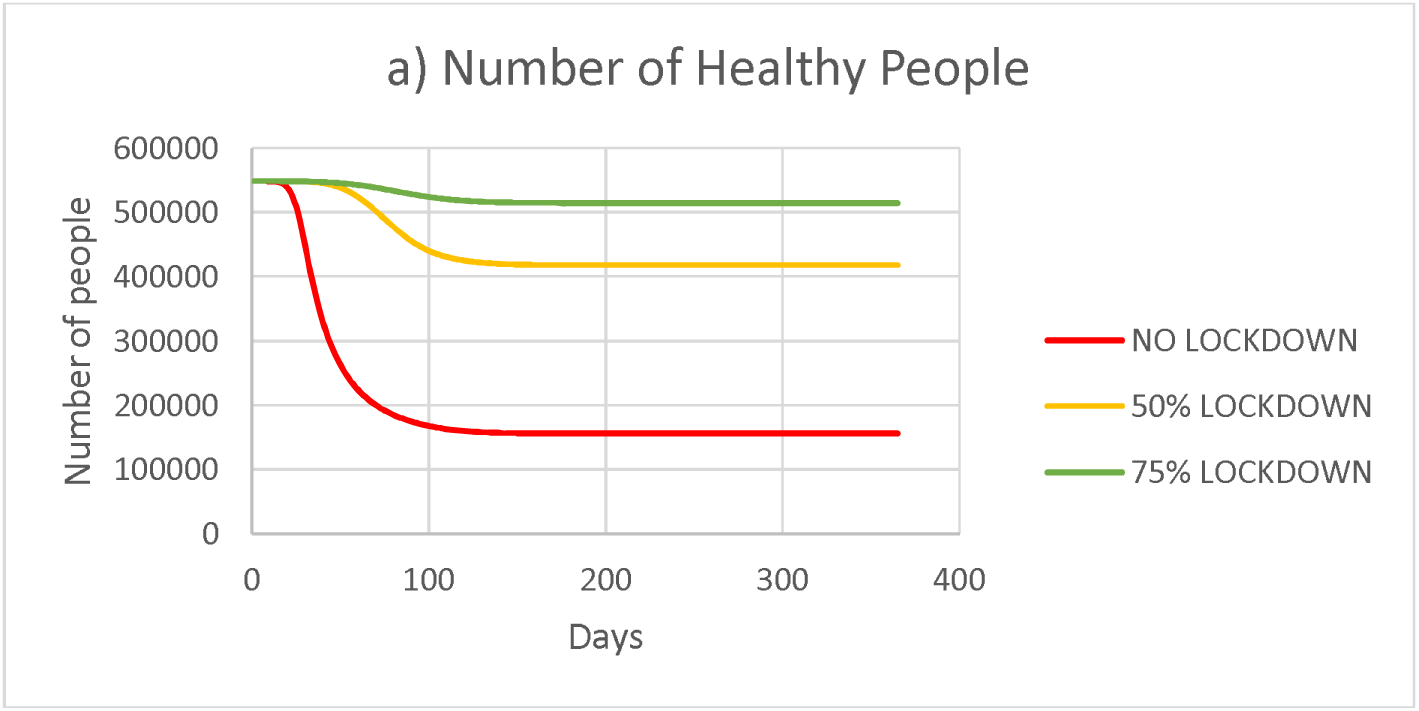
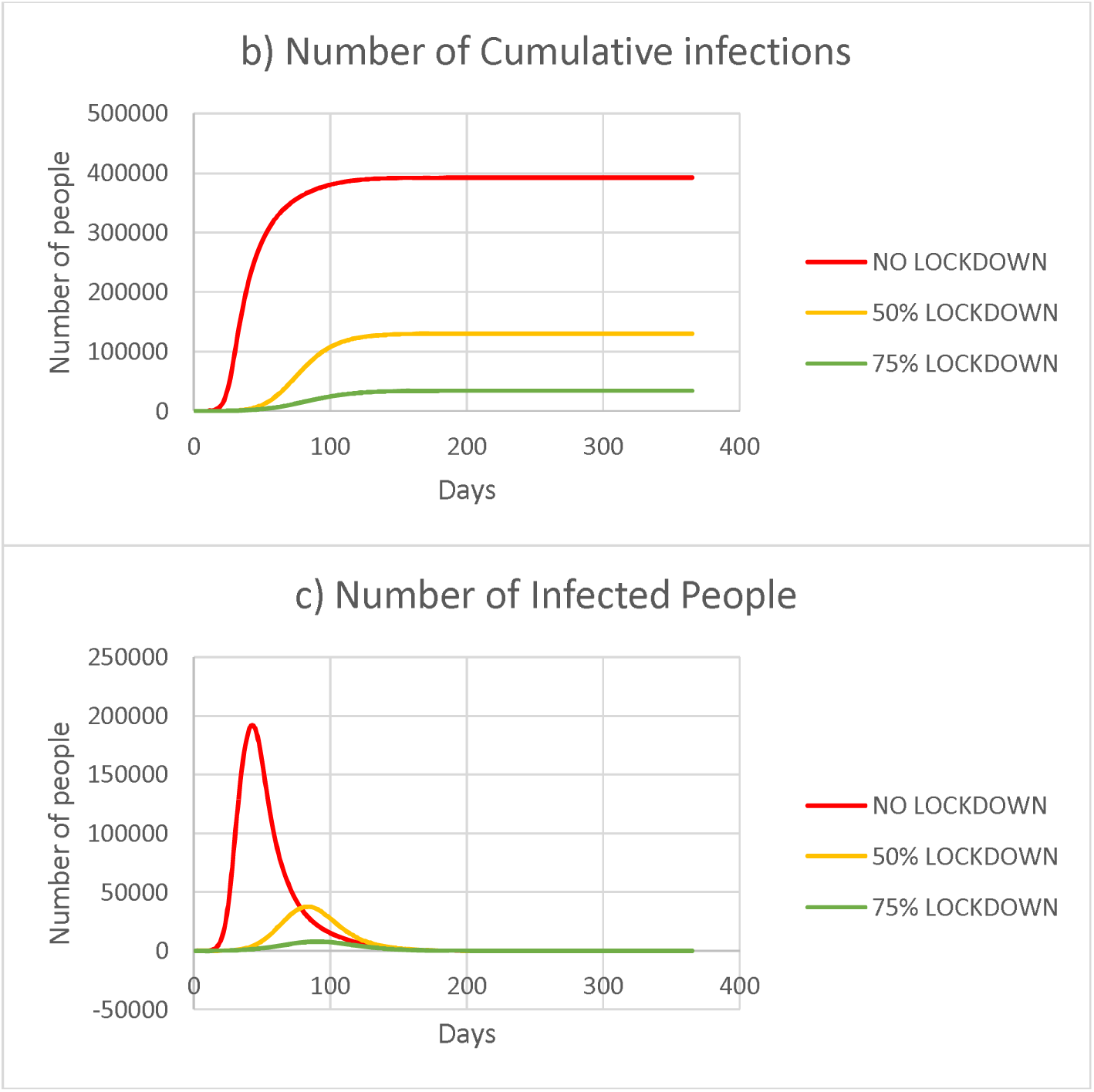
a) Number of uninfected people – all age groups b) Number of infected people – all age groups (Cumulative) c) Number of infected people (for a given instant)

The peak values in figure 3 a) corresponds to 31.71%, 6.28% and 1.33% of the initial uninfected population respectively for the three scenarios. There is an occurrence of a maximum equal to 173892 on 44^th^ day, 34414 on 84^th^ day and 7281 on 90^th^ day for the no lockdown, 50% and 75% lockdown scenarios respectively. The peak admissions correspond to 6.42%, 1.33% and 0.28% of the initial healthy population respectively for the three scenarios. The peak number of patients in ICU, 2390, 496 and 94 respectively for the three scenarios, which indicate the minimum number of intensive care setups required for the three scenarios. Similarly, figure 3 d) indicates the minimum number of ventilator setups, 1929, 405 and 78 for the three scenarios respectively.

**Figure 3:**
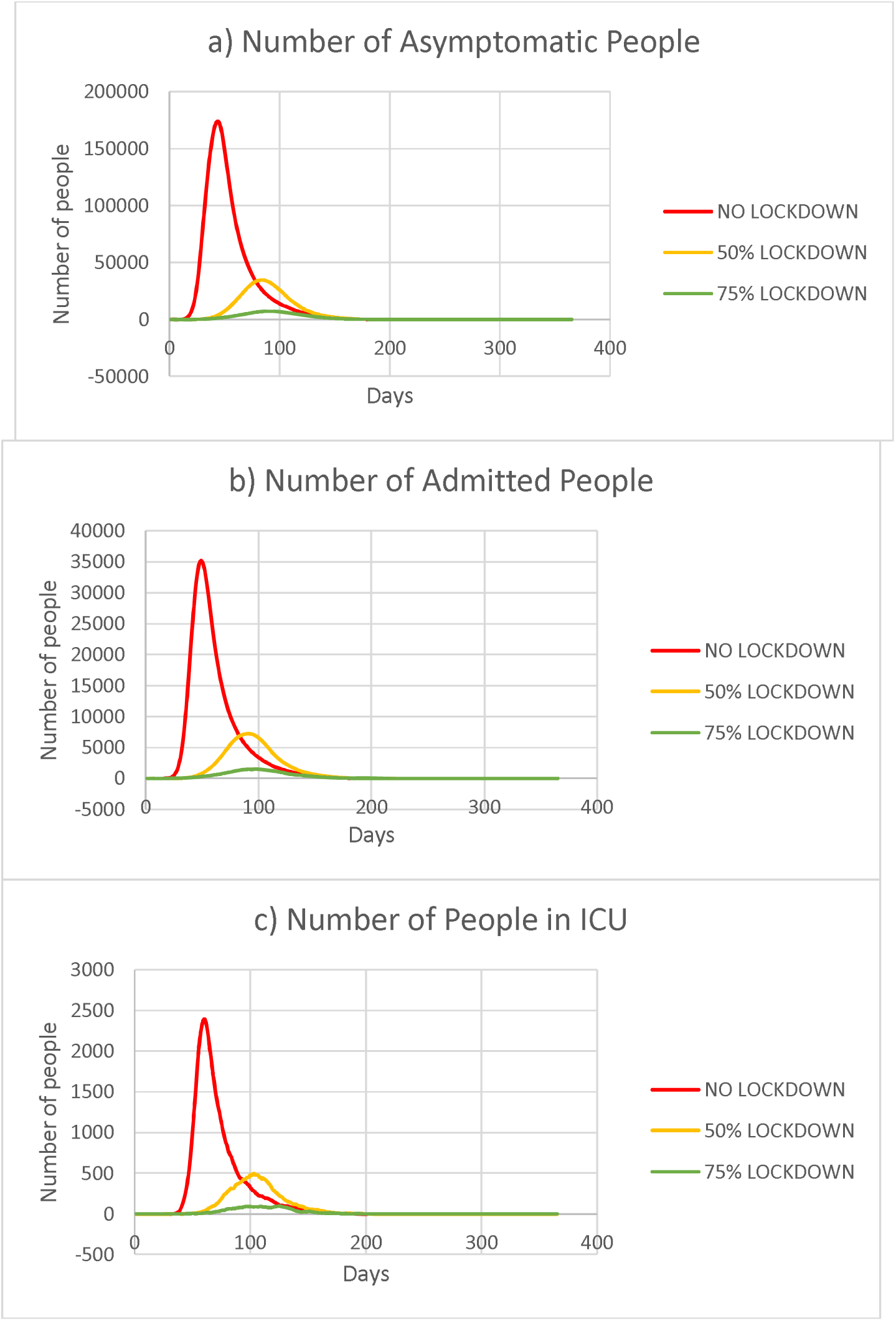
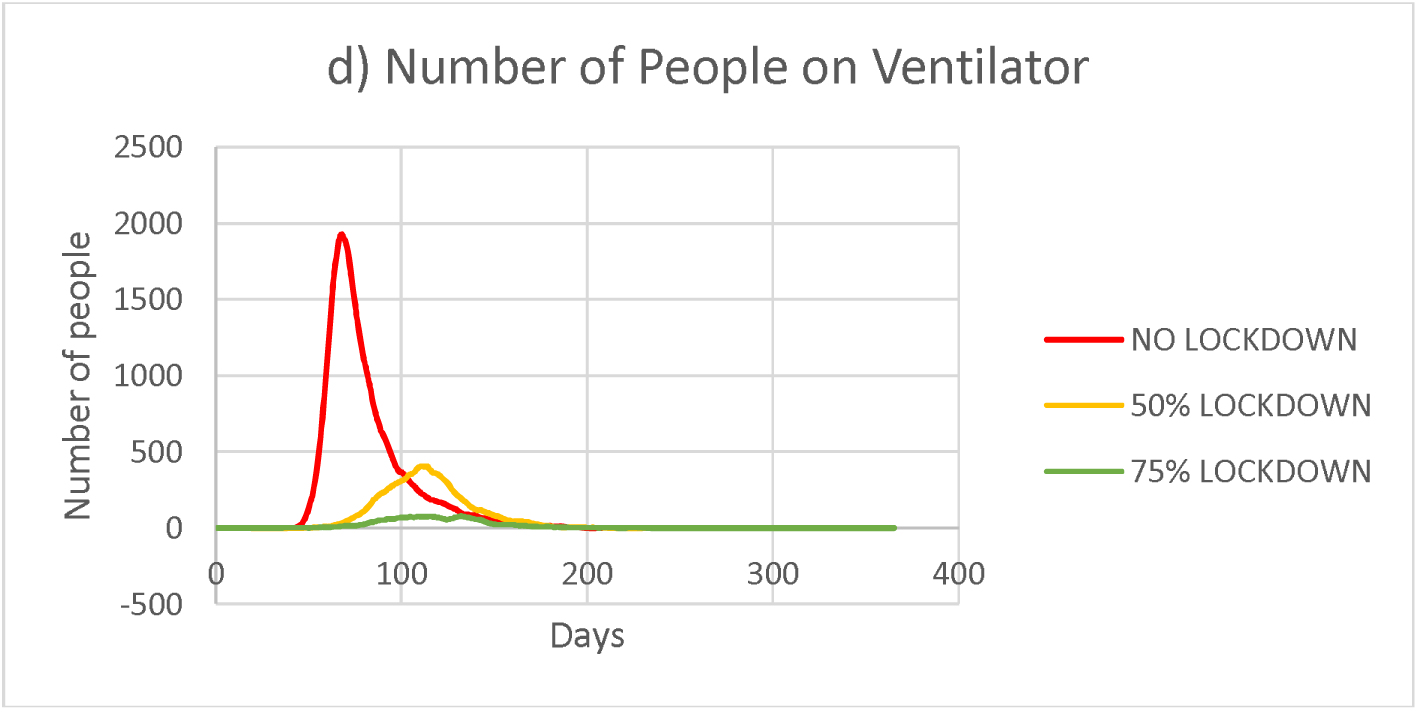
a) Number of asymptomatic people – all age groups b) Number of admitted people– all age groups c) Number of people in ICU – all age groups d) Number of people on ventilators – all age groups

Maximum deaths as seen in figure 4 correspond to 0.42%, 0.14% and 0.04% of the initial uninfected population respectively for three of the scenarios.

**Figure 4:**
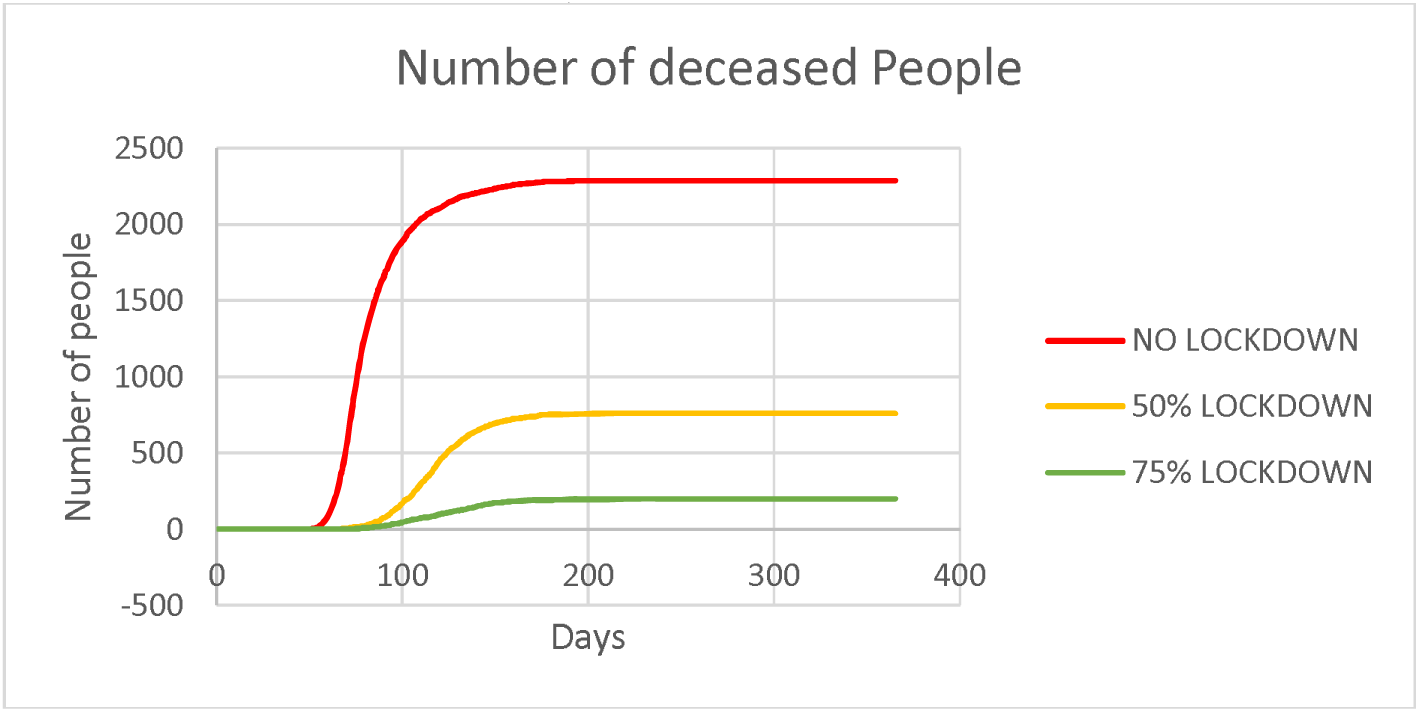
Number of people deceased – all age groups

**Figure 5:**
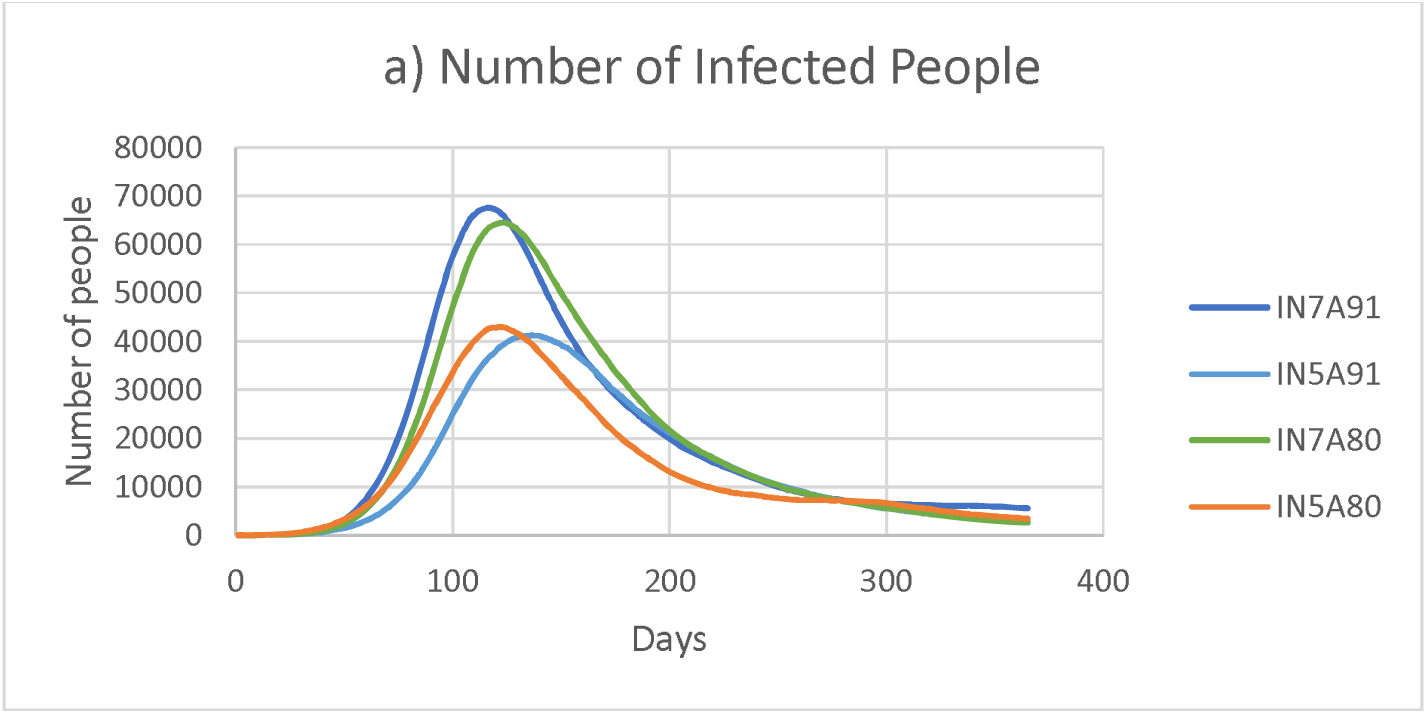
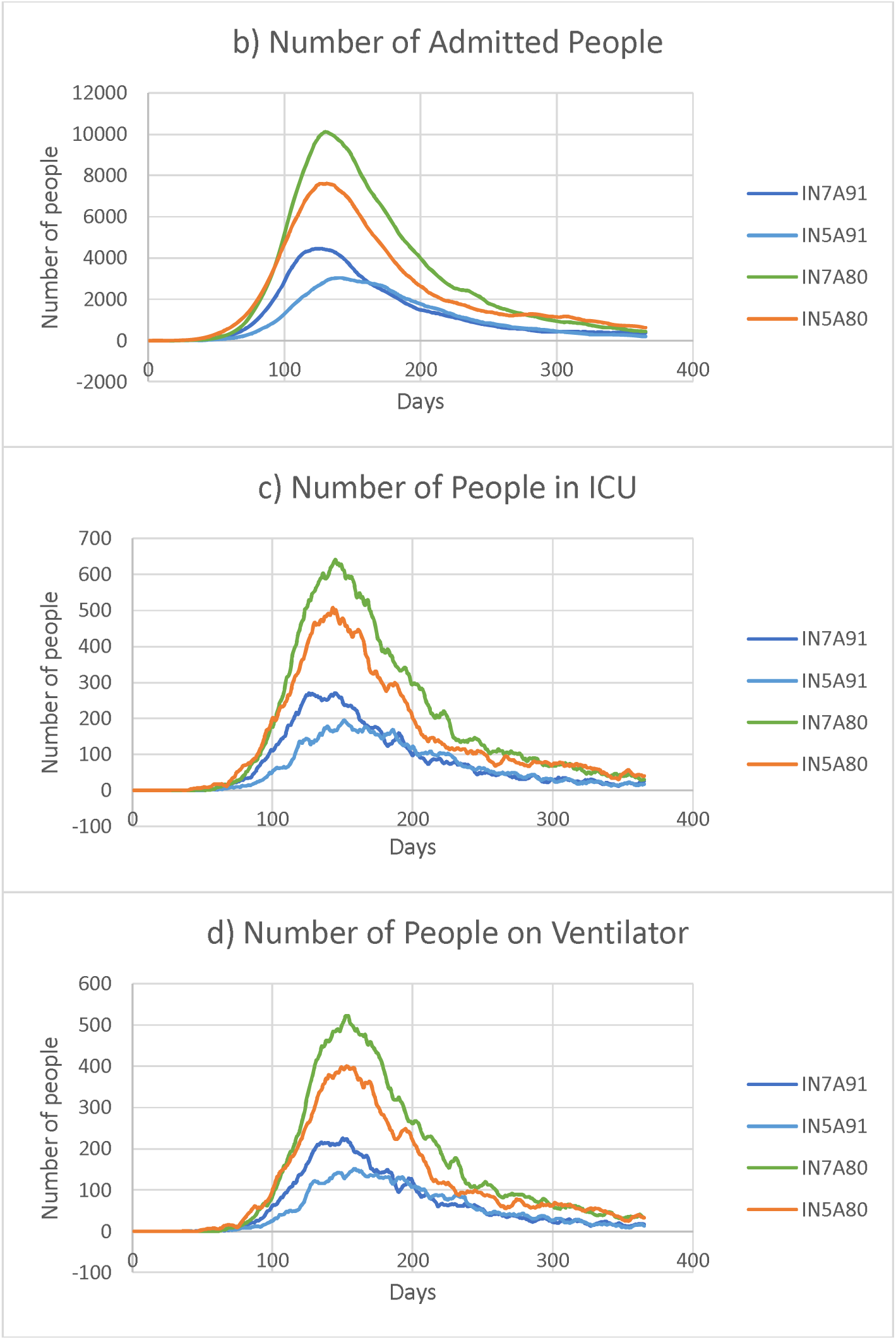
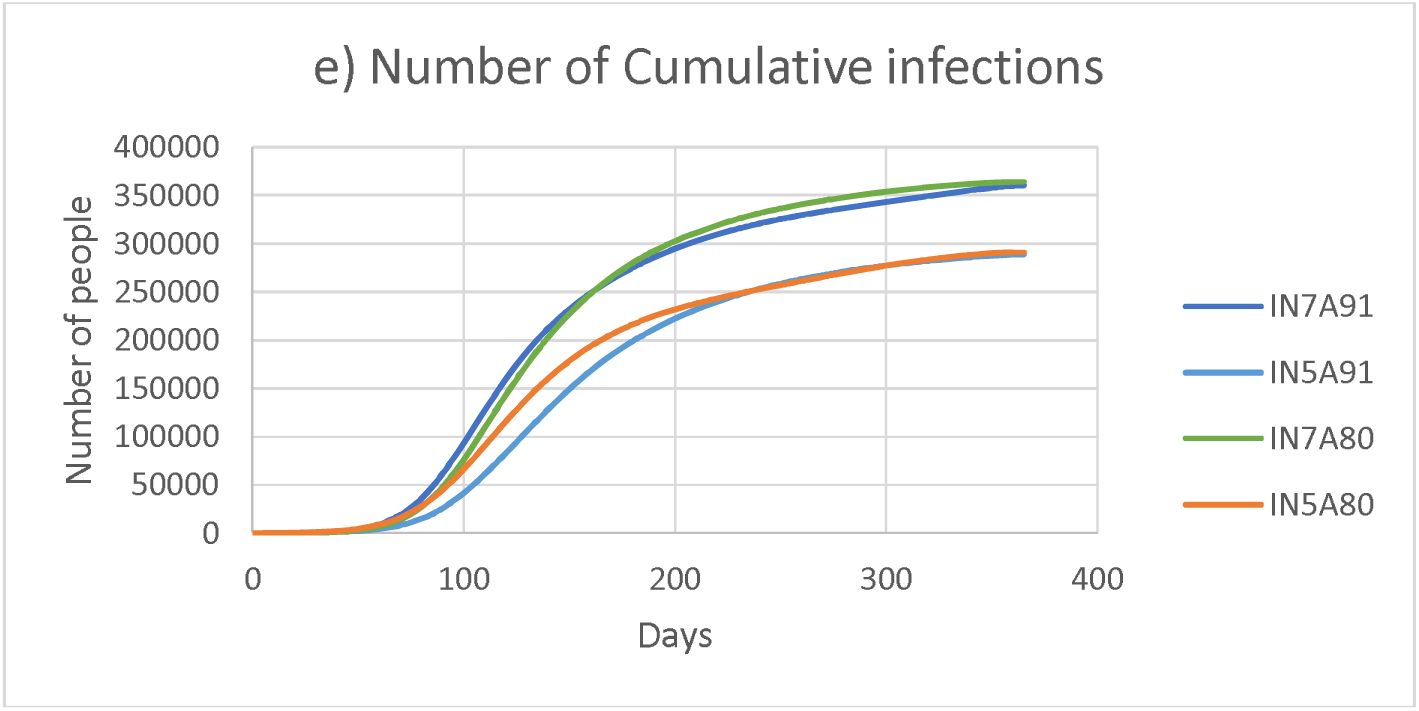
a) Number of Infected People, b) Number of Admitted People, c) Number of People in ICU, d) Number of People on Ventilator, e) Number of Cumulative infections

Table 4 indicates the peak values for various states possessed by the agents. The peak values also decrease for all these states as the stringency of lockdown is increased, indicating the effectiveness of lockdown measures. The values (in %) indicate the percentage with respect to the initial healthy population. A significant drop in peak number of ICUs required from 2390 for a no lockdown condition to 94 for a 75% lockdown is evident. Concurrently, the peak number of ventilators decline from 1929 for a no lockdown condition to 78 for a 75% lockdown condition.

**Table 4:** Peak values for various health conditions

|  | No lockdown |  |  | 50% lockdown |  |  | 75% lockdown |  |  |
| --- | --- | --- | --- | --- | --- | --- | --- | --- | --- |
|  | Value | % | Day | Value | % | Day | Value | % | Day |
| Uninfected | 548320 | 100 | 1 | 548321 | 100 | 1 | 548321 | 100 | 1 |
| Infected | 191907 | 35.00 | 43 | 37790 | 6.89 | 84 | 7986 | 1.46 | 90 |
| Asymptomatic | 173892 | 31.71 | 44 | 34414 | 6.28 | 84 | 7281 | 1.33 | 90 |
| Symptomatic | 23269 | 4.24 | 36 | 3450 | 0.63 | 78 | 723 | 0.13 | 85 |
| Admitted | 35186 | 6.42 | 49 | 7271 | 1.33 | 91 | 1510 | 0.28 | 97 |
| ICU | 2390 | 0.44 | 60 | 496 | 0.09 | 104 | 94 | 0.02 | 125 |
| Ventilator | 1929 | 0.35 | 68 | 405 | 0.07 | 114 | 78 | 0.01 | 132 |
| Deceased | 2288 | 0.42 | 224 | 760 | 0.14 | 209 | 199 | 0.04 | 231 |
| Recovered | 389572 | 71.05 | 226 | 129018 | 23.5 | 216 | 33773 | 6.16 | 230 |

## 4. Sensitivity Analysis

The uncertainties about the nature of COVID-19 has posted a great challenge to the healthcare fraternity. Yet, theories such as Swiss Cheese model that believes in partial curtailing of the spread through multiple strategies prove effective [29; 56]. The combined effect of multiple intervention scenarios such as closure of schools, work places, imposing lockdowns, use of control measures [29] and setting up of multiple defense mechanisms in healthcare facilities are in practice [56]. Recent studies in India showed longer means for incubation period (6.93 days) [59] and higher proportion of asymptomatic people (91%) [45]. Sensitivity analysis has been carried out by varying the asymptomatic proportion of infection and incubation period. A total of four scenarios have been compared using two different combinations of incubation period and asymptomatic proportion (Table 5).

**Table 5:** Parameters for Sensitivity Analysis scenarios and results

| Scenario | IN5A80 | IN5A91 | IN7A80 | IN7A91 |
| --- | --- | --- | --- | --- |
| Incubation period (days) | 5 | 5 | 7 | 7 |
| Asymptomatic proportion (%) | 80 | 91 | 80 | 91 |
| Asymptomatic | 37971 | 39297 | 56610 | 64091 |
| Symptomatic | 5169 | 2063 | 8165 | 3585 |
| Infected | 43066 | 41242 | 64526 | 67631 |
| Admitted | 7621 | 3045 | 10122 | 4466 |
| ICU | 508 | 196 | 640 | 270 |
| Ventilator | 400 | 151 | 523 | 225 |
| Peak Values Deceased | 611 | 289 | 767 | 327 |
| Cumulative Infected | 291326 | 288634 | 364270 | 360795 |

It is clear from the results of sensitivity analysis that the total infections are higher during longer incubation periods as people are assumed to spread the infection until they are either admitted or recovered. Just an increase of 2 days results in 25.03% and 25.11% increase in overall infections for scenarios with incubation period of 5 and 7 days respectively. Higher number of asymptomatic people (91%) increases demands increase of testing as they remain to be carriers until they recover. Lower number of admissions in scenarios with 91% asymptomatic people is since the asymptomatic proportion are assumed to recover without getting admitted. These projections are conservative in terms of showing the variations in spread due to change in driving parameters.

## 5. Discussion

This approach based on the synthetic population of 5,43,823 agents in Rangareddy District, for three different NPI scenarios (no lockdown, 50% lockdown and 75% lockdown) projects that the transmission rate of COVID-19 could be effectively brought down by stringency of lockdown measures. The study was performed at the district-level is a major strength of the study as it facilitates decision-making easier to policymakers at specific regions [10]. The simulation results are presented using ISPOR-SMDM Modeling Good Research Practices and ethical good practice in modelling.

Synthetic populations are most often generated from open source data such as Open data from London Imperial College, etc., [36], US Census Bureau data, [35; 39], Australian Census data [66], etc. The geographical scope of the study governs the number of agents. For example, 10 million stochastic agents for the State of Delaware, US [39], a scaled-down simulation of New York with 10000 agents [69], synthetic population of NYC with 500,000 [35], 5000 agents in the premises of a University in Italy [20], 24 million agent representing Australia [66], 750,805 agents representing Urmia, Iran [50], etc. In this study, we have scaled-down the synthetic population of Rangareddy district to 10.35% and used for the ABM.

Contact networks augment variations in behavioral aspects of agents in each model. Classification of a group of people aged greater than 65 and/ or with underlying illness as obesity, chronic cardiac or respiratory illness, and diabetes [36], awareness level (that enhances protection), use of contact tracing mechanisms [41], schedule-based contacts with house members [35; 39; 69], close contacts [35], closed spaces such as office spaces, university [20; 69], indirect contact with suspended viral particles, public gatherings at café, gym, hospitals, transport, [39], touching contaminated surfaces, washing hands [19; 41; 69], etc., have all been modeled. Present study modeled the contacts of agents based on the contact rates derived based on population densities.

There are also a spectrum of scenarios analyzed by modelers with an aim to determine the ones that outperform others, such as the no lockdown scenario that is included in almost all studies to be used as a base for comparison, control measures such as face masks, physical distancing, shielding of vulnerable population [36], lockdowns for varying durations [36], reducing contacts in external settings whilst maintaining the close contacts constant [39], lifting lockdowns based on age-groups [35], and effect of contact tracing of symptomatic individuals [35; 69]. NPIs such as 50 and 75 percent lockdowns have been used as scenarios in the present study.

In the present study, a decrease in number of contacts at various locations such as in schools, works, etc., was incorporated to enact lockdown scenarios whilst maintaining the contacts made at house [74]. It is evident from the results of present study that as the percentage of lockdown imposed was increased, the magnitudes of peak infections reduced with a delay in their corresponding occurrences, which provides more time for the policymakers to increase their capacities to meet the influx of cases. A team of researchers from The Center For Disease Dynamics, Economics & Policy (CDDEP) and Princeton University using ABM estimated the state-wise capacity requirements to accommodate the influx of hospitalizations to help the policymakers to increase their capacities to match the influx based on estimates in India [40; 73].

Considering some other parameters such clustering in contact networks, especially in the context of spread of infections would provide more accurate results [21; 47; 64; 70]. GIS information and migration routes could be included to improve the projections in specific areas [27; 60]. Wearable devices could be integrated with mobiles to provide real-time monitoring of COVID-19 patients [55]. Exploring the contact network and dynamics of different regions would help us to represent the region-specific disease spread better [10]

There are certain limitations to the study as parameters such as underlying health conditions, migration routes, adoption of control measures (face mask, social distancing, etc.), longitudinally varying lockdown phases, etc., have not been considered. The parameters which were used in the model were form different countries and may not represent the India or Rangareddy district scenario. The results of simulation model clearly indicate that the peak values could significantly be reduced by increasing the lockdown imposed. Thus, the importance of reducing the number of contacts, i.e., social distancing, is apparent through the results of this study and flattening the disease curve.

## 6. Conclusions

Majority of the ABM studies focus on specific regions that is a major strength of ABM as it allows defining characteristics at individual level [35]. We present this ABM using AnyLogic on a synthetic population in Rangareddy district, Telangana state, India. Further, data specific to India to parametrize such ABM will be critical. Having a synthetic population of a country can provide several options to create ABMs for several disease conditions apart from COVID-19 and may prove efficient for decision-making.

## Data Availability

Detailed day-wise data of the number of people in each of the state (age-group wise), the AnyLogic model file, synthetic population (text file) and input template (Spreadsheet) are available in the links provided.

https://cloud.anylogic.com/model/7cd10c0c-f1c1-4b8f-9aac-0bf37a45379a?mode=SETTINGS

https://osf.io/utmhg/?view_only=05ac26fc100645be8b1bba6557d606be

## Data Availability

Detailed output (age-wise), AnyLogic model file, synthetic population and input spreadsheet are available at: https://cloud.anylogic.com/model/7cd10c0c-f1c1-4b8f-9aac-0bf37a45379a?mode=SETTINGS and https://osf.io/utmhg/?view_only=05ac26fc100645be8b1bba6557d606be

